# Identifying Individuals at High Risk for Alzheimer’s Disease Among Hispanics Using Single and Multi-Ancestry Polygenic Risk Scores

**DOI:** 10.1101/2024.10.21.24315877

**Authors:** Yuexuan Xu, Min Qiao, Tamil I. Gunasekaran, Yan Gu, Dolly Reyes-Dumeyer, Angel Piriz, Danurys Sanchez, Belisa Soriano, Yahaira Franco, Zoraida Dominguez Coronado, Patricia Recio, Diones Rivera Mejia, Martin Medrano, Rafael A. Lantigua, Lawrence Honig, Jennifer J. Manly, Adam M. Brickman, Badri N. Vardarajan, Richard Mayeux

## Abstract

**Introduction:** Polygenic risk score (PRS) is effective in predicting AD risk among Europeans but remains understudied in Hispanics. Diverse genome-wide association studies (GWAS) data across multiple ancestries may improve PRS predictions. We evaluated PRS performance to predict AD disease risk using novel methods in the largest available African, European, and Hispanic GWAS for AD.

**Methods:** Prediction performance of *APOE*, single-ancestry PRS, and multi-ancestry PRS derived from GWAS-focused and method-focused approaches to clinical AD, incident AD, and cognition were evaluated in 2,961 Hispanics from two large studies. The GWAS-focused approach constructs PRS based on multi-ancestry GWAS, while the method-focused approach uses novel multi-ancestry PRS methods, integrating GWAS summary statistics across ancestries. Ten repetitions of 5-fold cross-validation were used. In a subset, plasma biomarker data were used in a tuning-validation split to examine PRS performance in predicting single and combined biomarkers.

**Findings:** The multi-ancestry PRS excluding *APOE,* constructed using the method-focused approach, outperformed both single-ancestry and multi-ancestry PRSs from the GWAS-focused approach. The best method-focused PRS, incorporating summary statistics from GWASs of African, European, and Hispanic populations, explained up to 1.6%, 3.9%, and 1.7% of the variance in clinical AD, incident AD, and cognition, respectively - comparable to or even higher than the variance explained by the *APOE*. Similar findings were observed in biomarker analyses. *APOE* accounted for more variation in plasma P-tau levels and PRS explained more variation in Aβ levels.

**Interpretation:** Integrating novel multi-ancestry PRS methods with GWAS across ancestries enhances prediction accuracy for AD risk among Hispanics. *APOE* and PRS may point to different biological aspects of AD.

**Funding:** National Institutes of Health R01 AG072474, RF1 AG066107, 5R37AG015473, RF1AG015473, R56AG051876, R01 AG067501, and UL1TR001873.

**Research in context:** *Evidence before this study:* We searched PubMed for research related to PRS prediction of AD in Europeans, Africans, and Hispanics, published from database inception to August 1, 2024, without any language restrictions. The search terms used were "PRS," "PGS," "Hispanics," "Latinos," "AD," and "plasma biomarkers." We considered only peer-reviewed reports in English. Previously, few studies have examined the performance of PRS in predicting clinical AD, incident AD, and mild cognitive impairment (MCI) among Hispanics. However, none of these studies utilized advanced methods for constructing multi-ancestry PRS, validated PRS performance among Hispanics, or examined plasma biomarkers.

*Added value of this study:* The present study demonstrated that integrating novel multi-ancestry PRS methods with GWAS from African, European, and Hispanic populations enhanced prediction accuracy for AD risk among Hispanics. Among Hispanics, PRS explains a similar or higher amount of variance compared to *APOE*. Plasma biomarker analyses suggests that *APOE* may also be strongly related to variation in P-tau levels, while PRS may explain variance in Aβ levels.

*Implications of all the available evidence:* Among Hispanics, PRS complements the effects of *APOE* and has the potential to identify at-risk populations for clinical trial eligibility and early biomarker screening. Although AD genetic studies are still limited among Hispanics, a dynamic combination of advanced methods with GWAS across populations could substantially improve prediction performance in this population, which in turn may reduce health disparities.

## Introduction

Alzheimer’s disease (AD) is a progressive brain disorder with high heritability, influenced by common and rare genetic variants. The polygenic risk score (PRS), which measures genetic predisposition to AD, is effective for risk stratification, achieving up to 84% area under the curve for predicting AD in European populations^1^.

Hispanics have a higher risk for AD than non-Hispanic whites^2^, but the effect of *APOE*, the strongest genetic risk factor for AD, is less pronounced in this group^3^. This observation suggests that other genetic variants may contribute to AD risk in Hispanics, highlighting the need for PRS-based risk assessment. Hispanics are underrepresented in PRS research due to the lack of Hispanic-specific genome-wide association studies (GWAS) and limited inclusion in population-based AD studies. A few studies explored PRS performance in Hispanics; however, these studies either used GWAS data from non-Hispanic populations or summary statistics from exploratory Hispanic GWAS, with limited statistical power and applied single-ancestry PRS methods that need further validation^4–6^. Researchers have also conducted meta-analyses of GWAS in AD across diverse populations and developed a multi-ancestry PRS for a three-way admixed Colombian population^7^. However, the prediction performance of this PRS remains limited, especially when excluding *APOE*.

Optimal PRS predictive power may be achieved by using GWAS across multiple ancestries for prediction within any single ancestry^8^. Building on this concept, several advanced trans-ancestry PRS methods have been developed to enhance prediction in underrepresented populations. For example, one Bayesian method used multivariate priors for effect-size distribution to leverage information across populations^9^. Improved PRS accuracy in underrepresented populations may also be achieved by harnessing shared genetic effects across ancestries^8^. Additionally, some studies integrated machine-learning techniques (e.g., ensemble learning) with empirical Bayes methods, clumping and thresholding approaches, or combinations of lasso and ridge penalty functions to further boost PRS performance across diverse populations^10,11^. However, a thorough examination of the prediction performance of these advanced PRS methods for AD among Hispanics is warranted.

In this study, we assessed PRS performance among Hispanics by developing single-ancetsry and multi-ancestry PRS using advanced trans-ancestry PRS methods, based on data from the Estudio Familiar de Influencia Genética en Alzheimer (EFIGA) and the Washington Heights-Hamilton Heights-Inwood Columbia Aging Project (WHICAP).

## Method

An overview of all methods is provided in eMethods. This genetic association study followed the STREGA guideline, with written informed consent obtained from participants prior to participation. The study protocols for EFIGA and WHICAP were approved by the Columbia University Medical Center IRB, and the National Council of Bioethics in Health (CONABIOS) of the Dominican Republic approved the study protocol for EFIGA.

### Data, ascertainment of genotype and phenotype

The data analyzed in this study came from individuals of Caribbean Hispanic ancestry enrolled in either the EFIGA or WHICAP study^12,13^. We combined data from both studies to assess PRS performance. Although PRS derived from Hispanic-specific GWAS was of particular interest, no external Hispanic GWAS was available. Therefore, we leveraged summary statistics from an internal GWAS of 5,110 Hispanics, previously published^5^. To avoid overfitting, we excluded participants from the internal GWAS from the final PRS assessment, leaving a sample of 2,961 unrelated Hispanics.

PRS prediction performance was evaluated for three AD and related outcomes: clinically diagnosed AD, cognitive test performance, and AD-related plasma biomarker concentrations. EFIGA and WHICAP provided dementia status using the same diagnostic criteria and protocols. All AD diagnoses met the National Institute on Aging-Alzheimer’s Association criteria (McKhann criteria)^14^, while cognitively normal subjects had no AD or mild cognitive impairment (MCI) diagnosis.

All participants completed a neuropsychological battery assessing memory, language, visuospatial ability, and processing speed. The memory composite score, derived from factor analysis, was used as the main cognitive outcome due to its importance in AD and fewer missing data^15,16^.

We also analyzed AD-related plasma biomarker concentrations in a subset of participants, including plasma beta-amyloid (Aβ)42, Aβ42/40 ratio, phosphorylated tau181 (P-tau181), and P-tau181/Aβ42 ratio. Details on plasma acquisition and processing are described elsewhere^15^. Biomarker values were log-transformed due to skewed distributions. As studies suggest that combined plasma biomarkers offer better predictive value than individual ones, we conducted principal component analyses (PCAs) on four biomarkers—Aβ42/40, P-tau181/Aβ42, neurofilament light (NfL), and glial fibrillary acidic protein (GFAP) — and used the first two principal components (PCs) as indicators of combined biomarkers, as previously described^17^. NfL, GFAP, and P-tau181/Aβ42 were the primary contributors to PC1 (loadings ranging from 0.65 to 0.82), all exhibiting positive loadings. For PC2, Aβ42/40 and P-tau181/Aβ42 were the dominant biomarkers with the largest loadings, with Aβ42/40 having the largest positive loading (0.87) and P-tau181/Aβ42 a negative loading (-0.55). Based on the loading direction, a higher PC1 score suggests an increased likelihood of neuronal injury, neuroinflammation, and neurodegenerative profiles, while a higher PC2 score indicates a lower likelihood of AD-specific pathological changes.

Genetic data underwent standard quality control, imputation to the Trans-Omics for Precision Medicine reference panel (array-based samples), and ancestry determination. We used the PC-relate method to refine ancestry while adjusting for familial relatedness, and PC-AiR to account for population stratification.

### Polygenic risk score and APOE

Caribbean Hispanics represent an admixed population; therefore we leveraged the largest available AD GWAS summary statistics from European, African, and Hispanic ancestries to construct both single-ancestry and multi-ancestry PRS using various methods (Table 1, Supplementary Table 1)^5,18,19^. For single-ancetsry PRS, we employed three methods: clumping and thresholding (C+T), Lassosum2^20^, and PRS-CS^21^. For multi-ancestry PRS, we constructed both GWAS-focused and method-focused PRS.

**Table 1.** Summary of Methods Used in PRS Development.

| Method | # Source Populations | Tuning sample? | Description | Tuning parameters |
| --- | --- | --- | --- | --- |
| ***Single-ancestry PRS methods*** |  |  |  |  |
| C + T | single | Yes | a) LD-based clumping<br>b) *P*-value thresholding | *P*-value threshold |
| Lassosum2 | single | Yes | a) Lasso regression-based | $\delta$ and $\lambda$ combinations |
| PRS-cs | single | Yes | a) Bayesian shrinkage | Global shrinkage parameters |
| ***'Methods'-focused multi-ancestry PRS methods*** |  |  |  |  |
| BridgePRS | multiple | Yes | a) Fits Bayesian ridge regression to auxiliary GWAS<br>b) Using the posterior SNP effect sizes as priors for a second Bayesian ridge regression on the target non-European population. | $\lambda, \tau, \alpha, k$ |
| CTSLEB | multiple | Yes | a) C + T<br>b) Empirical-Bayes<br>c) Super learning | $r^2$-cut off and base size of the clumping window size, and *P*-value threshold |
| PRS-CSx | multiple | Yes | a) Models GWAS and LD across populations using a Bayesian framework with continuous shrinkage prior<br>b) Linearly combine the posterior effect-size estimates across populations using weights derived from a simple linear regression in the target population's tuning samples. | Global shrinkage parameters, Linear weights |
| PRS-CSx-meta | multiple | No | a) Models GWAS and LD across populations using a Bayesian framework with continuous shrinkage prior<br>b) Combine SNP effect sizes across populations with inverse-variance-weighted meta-analysis of population-specific posterior estimates. | - |
| PROSPER | multiple | Yes | a) Combines LASSO (L1) and Ridge (L2) penalties to regularize SNP effect sizes.<br>b) An ensemble step combines PRS scores generated across different penalty parameters and populations. | $\delta, \lambda, c_1, c_2, i, i_1, i_2 = 1, c \dots M$ |
| ***GWAS-focused multi-ancestry PRS methods*** |  |  |  |  |
| C + T | multiple | Yes | a) LD-based clumping<br>b) *P*-value thresholding<br>c) Based on multi-ancestry GWAS | *P*-value threshold |
| PRS sum | multiple | Yes | d) LD-based clumping<br>e) *P*-value thresholding<br>f) Simple sum of single-ancestry PRS | *P*-value threshold |
| Weighted PRS | multiple | Yes | a) LD-based clumping<br>b) *P*-value thresholding<br>c) Linear combination of single-ancestry PRS | *P*-value threshold, Linear weights |

The GWAS-focused multi-ancestry PRS was derived from a published multi-ancestry GWAS that includes European, Asian, African, and Hispanic populations^7^. Additionally, we created PRSs from in-house meta-analyses combining different population groups (European+African, European+Hispanic, and European+African+Hispanic), and used only the C+T method for these due to the difficulty in defining an LD reference panel that matches the multi-ancestry GWAS populations, which is essential for applying advanced methods, such as PRS-CS^21^. We also constructed multi-ancestry PRS using simple/weighted sums of single-ancestry PRS^6^.

For methods-focused multi-ancestry PRS, we first calculated a PRS incorporating large European GWASs and a smaller non-European GWAS, using a combination of C+T, the empirical-Bayes method, and a super learning model (CTSLEB)^11^. We also applied the novel two-stage Bayesian ridge method that leverages shared genetic effects across ancestries (BridgePRS)^8^. Furthermore, we calculated a multi-ancestry PRS integrating GWASs from more than two populations using posterior effect sizes inferred under coupled continuous shrinkage priors across populations (PRS-CSx/PRS-CSx-meta) and a combination of lasso and ridge penalty functions with ensemble learning to enhance PRS prediction performance across diverse populations (PROSPER)^9,10^. All methods except PRS-CSx-meta required a tuning cohort for parameter optimization. Several methods also required specifying a reference panel to infer the expected correlation structure; we used the LD populations recommended by each software implementation to construct a method-focused multi-ancestry PRS. For each GWAS-focused multi-ancestry PRS, we used our target sample as the reference panel. For each single-ancestry PRS, we used reference panels from the 1000 Genomes Project that match the ancestry of the population used for the corresponding GWAS, as previously recommended^22^. Default or recommended parameters/algorithms were used for all methods. The *APOE* region (GRCh38, chr19:43,907,927–45,908,821) was excluded from PRSs due to the better predictive performance of the *APOE*(ε2 + ε4) variant^23^. Lacking beta estimates for *APOE* polymorphisms rs7412 and rs429358 in European and Hispanic GWASs, we therefore used binary *APOE*-ε4 carrier status and the ancestry-specific *APOE* effect from the largest overview to construct an ancestry-specific-*APOE* score^3,24^.

### Statistical analyses and internal validation

We used two internal validation strategies to evaluate PRS performance with clinical AD, cognition, and plasma biomarkers. First, for AD and cognition, we performed ten repeated 5-fold cross-validations to ensure robust performance estimates and minimize overfitting, particularly for methods involving machine learning. This approach was chosen for computational efficiency, as minimal improvement is seen beyond 10 repetitions^25^. Prediction accuracy and variability were assessed using pooled-effect sizes through the inverse-variance method (odds ratio [OR], hazard ratio [HR], or beta), mean area under the curve (AUC, for AD status), and mean incremental R² (Δ*R^2^*) with 95% confidence intervals. Differences between methods were tested using a corrected pair-wise t-test for cross-validated results^26^. For methods that do not require parameter tuning, we reported the average performance across validation datasets.

Second, for biomarker analyses, because plasma biomarker concentrations were available only in a subset of participants, we used a tuning-validation split. Parameter tuning was conducted among participants without biomarker data, and performance was validated with those who had biomarker data. To assess the PRS’s effectiveness in predicting AD status, we used logistic regression, and to predict cognition and biomarker concentrations, we used linear regression, with age, sex, cohort, and the first 10 PCs as covariates. We also used Cox proportional hazards analysis to evaluate PRS performance in predicting conversion from cognitively normal to AD. This analysis included participants with at least one year of follow-up, retaining only incident cases and controls, with age, sex, cohort, and the first 10 PCs as covariates. A two-sided *P*-value<0.05 was considered statistically significant. For all analyses, continuous PRS and the *APOE* indicator were z-standardized. All statistical analyses were conducted in R v4.2.2.10.

## Results

### APOE and single-ancestry PRS

The sample sizes for AD, incident AD, cognition, and biomarkers were 2,961, 1,094, 2,646, and 631, respectively (Table 2). The mean age ranged from 72 to 75 years, with 70% women, an average education of 6 years, and 30% *APOE*-ε4 carriers. AD cases accounted for 30%-40% of the sample across outcomes. No differences in variable distribution were observed between tuning and validation datasets in repeated cross-validation (results available upon request).

**Table 2.** Descriptive Statistics of the Study Sample Across Different Outcomes.

|  | **AD status, N = 2,961<sup>†</sup>** | **Incident AD, N = 1,094<sup>†</sup>** | **Cognition, N = 2,646<sup>†</sup>** | **Biomarkers, N = 631<sup>†</sup>** |
| --- | --- | --- | --- | --- |
| Age (years) | 74 (9) | 72 (7) | 73 (9) | 75 (9) |
| Sex |  |  |  |  |
| Men | 902 (30%) | 321 (29%) | 814 (31%) | 180 (29%) |
| Women | 2,059 (70%) | 773 (71%) | 1,832 (69%) | 451 (71%) |
| Education (years) | 6 (5) | 7 (5) | 6 (5) | 6 (5) |
| APOE-ε4 |  |  |  |  |
| No | 2,012 (68%) | 784 (72%) | 1,810 (68%) | 430 (68%) |
| Yes | 949 (32%) | 310 (28%) | 836 (32%) | 201 (32%) |
| AD |  |  |  |  |
| Controls | 1,869 (63%) | 694 (63%) | 1,847 (70%) | 379 (60%) |
| Cases | 1,092 (37%) | 400 (37%) | 799 (30%) | 252 (40%) |
| Cohort |  |  |  |  |
| EFIGA | 2,148 (73%) | 514 (47%) | 1,984 (75%) | 429 (68%) |
| WHICAP* | 813 (27%) | 580 (53%) | 662 (25%) | 202 (32%) |

<sup>†</sup>Mean (SD); n (%)
\* This study focuses on Caribbean Hispanics from WHICAP, a community-based, multiethnic study of elderly individuals aged 65 and older, residing in northern Manhattan, New York City. While WHICAP includes non-Hispanic white and Black individuals, we limit the current analyses to Caribbean Hispanic participants.

*APOE*-ε4 consistently showed a strong association with clinical AD among Hispanics (pooled OR: 1.83 [1.72-1.94], mean ΔR² = 0.014 [0.011-0.016], mean AUC = 0.60 [0.59-0.62]), incident AD (pooled HR: 1.43 [1.34-1.54], mean ΔR² = 0.021 [0.016-0.027]), and cognition (pooled beta: -0.78 [-0.85 to -0.71], mean ΔR² = 0.015 [0.013-0.017]). The *APOE* score explained slightly more variance than the binary *APOE*-ε4 indicator, but performed similarly when accounting for *APOE* effects across ancestries (Figure 1).

**Figure 1.**
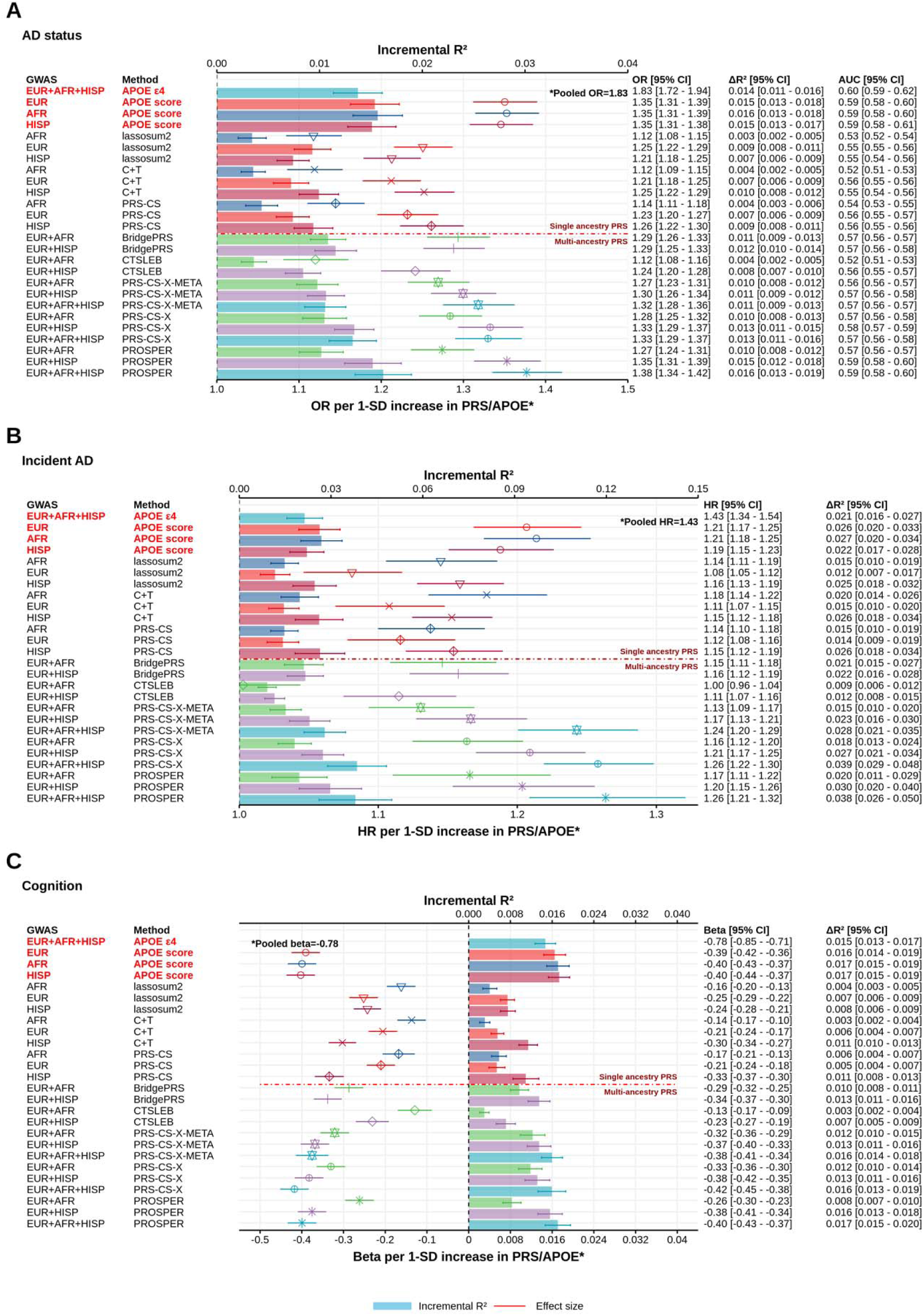
Performance of *APOE*, ancestry-specific, and ‘method’-focused multi-ancestry PRS in predicting clinical AD, incident AD, and cognition among Hispanics. Figure 1 presents the mean performance of *APOE*, single-ancestry, and multi-ancestry PRS derived from the method-focused multi-ancestry approach, integrating GWAS from African, European, and Hispanic populations, in predicting clinical AD (A), incident AD (B), and cognition (C). Within each panel, each point with an error bar represents the pooled effect size of the corresponding PRS based on 10-repeated 5-fold cross-validation with 95% confidence intervals. Each bar with an error bar represents the mean incremental R² for different approaches, also based on 10-repeated 5-fold cross-validation with 95% confidence intervals. The performance of each PRS within each fold and repeat for clinical AD, incident AD, and cognition was evaluated using logistic regression, Cox proportional hazards models, and linear models, respectively, with age, sex, cohort, and the first 10 principal components as covariates. The mean AUC was calculated as the confounder-adjusted AUC for the PRS, following 10-repeated 5-fold cross-validation. The *APOE* score for each ancestry was weighted by the corresponding *APOE* genotype effect size based on a recent large overview of *APOE* effects across ancestries. All continuous genetic predictors have been standardized to a mean of 0 and a standard deviation of 1 for ease of comparison.

Single-ancestry PRS generally explained 1% or less of the variance for clinical AD (pooled OR: 1.12 to 1.26), 1.2%-2.6% for incident AD (pooled HR: 1.08 to 1.18), and 0.3%-1.1% for cognition (pooled beta: -0.33 to -0.14). PRS from Hispanic GWASs generally performed as well or slightly better than those from European or African GWASs, except for the PRS derived from lassosum2 in predicting clinical AD (Figure 1).

### Method-focused multi-ancestry PRS

Figure 1 shows the average performance of PRS from method-focused multi-ancestry approaches for predicting clinical AD, incident AD, and cognition. For clinical AD, multi-ancestry PRS generally performed as well as or better than single-ancestry PRS. PRS based on all three GWASs generally performed as well or better than those based on two. In 10-fold cross-validation, PROSPER, integrating European, African, and Hispanic GWASs, provided the most predictive PRS on average (pooled OR: 1.38 [1.34-1.42], mean ΔR² = 0.016 [0.013-0.019], mean AUC = 0.59 [0.58-0.60]) (Figure 1A). This approach showed a significant improvement of 0.6%-1.3% in ΔR² and 0.036-0.07 in AUC over the 10-fold cross-validated single-ancestry PRS, regardless of the GWAS (Supplementary Figure 1). Smaller but similar improvements were observed for PRS-CSx and PRS-CSx-meta when integrating European, African, and Hispanic GWASs. When combining European GWAS with smaller African or Hispanic GWASs, the *European-Hispanic* PRS generally performed better than the *European-African* PRS, though the latter still performed as well as or better than most single-ancestry PRS, except those derived from CTSLEB.

Similar findings were observed for incident AD and cognition. PROSPER, integrating European, African, and Hispanic GWASs, provided the most predictive PRS on average for incident AD (pooled HR: 1.26 [1.21-1.32], mean ΔR² = 0.038 [0.026-0.050]) and cognition (pooled beta: -0.40 [-0.43 to -0.37], mean ΔR² = 0.017 [0.015-0.020]) (Figure 1B-1C). PRS constructed from GWASs across all three ancestries generally outperformed those from two, with PRSs from two ancestries often performing as well or better than most single-ancestry PRS. However, some methods integrating European with African or Hispanic GWASs showed improved performance, whereas others slightly underperformed compared to PRS derived from single-ancestry Hispanic GWAS, particularly in predicting incident AD (Supplementary Figure 2).

### GWAS-focused multi-ancestry PRS

PRS constructed from meta-analyses of GWASs across ancestries, or linear combinations of single-ancestry PRS generally explained less than 1% of the variance in clinical AD (pooled OR: 1.12 to 1.24), less than 2% for incident AD (pooled HR: 1.07 to 1.14), and less than 1% for cognition (pooled beta: -0.19 to -0.27) (Figure 2). PRS from meta-analyses of GWASs across ancestries performed as well as or worse than those from Hispanic or European GWASs across outcomes, regardless of the number of GWASs included, whether from in-house analyses or Lake et al^7^. Similarly, neither simple nor weighted combinations of single-ancestry PRS outperformed PRS from Hispanic GWAS (Supplementary Figure 3).

**Figure 2.**
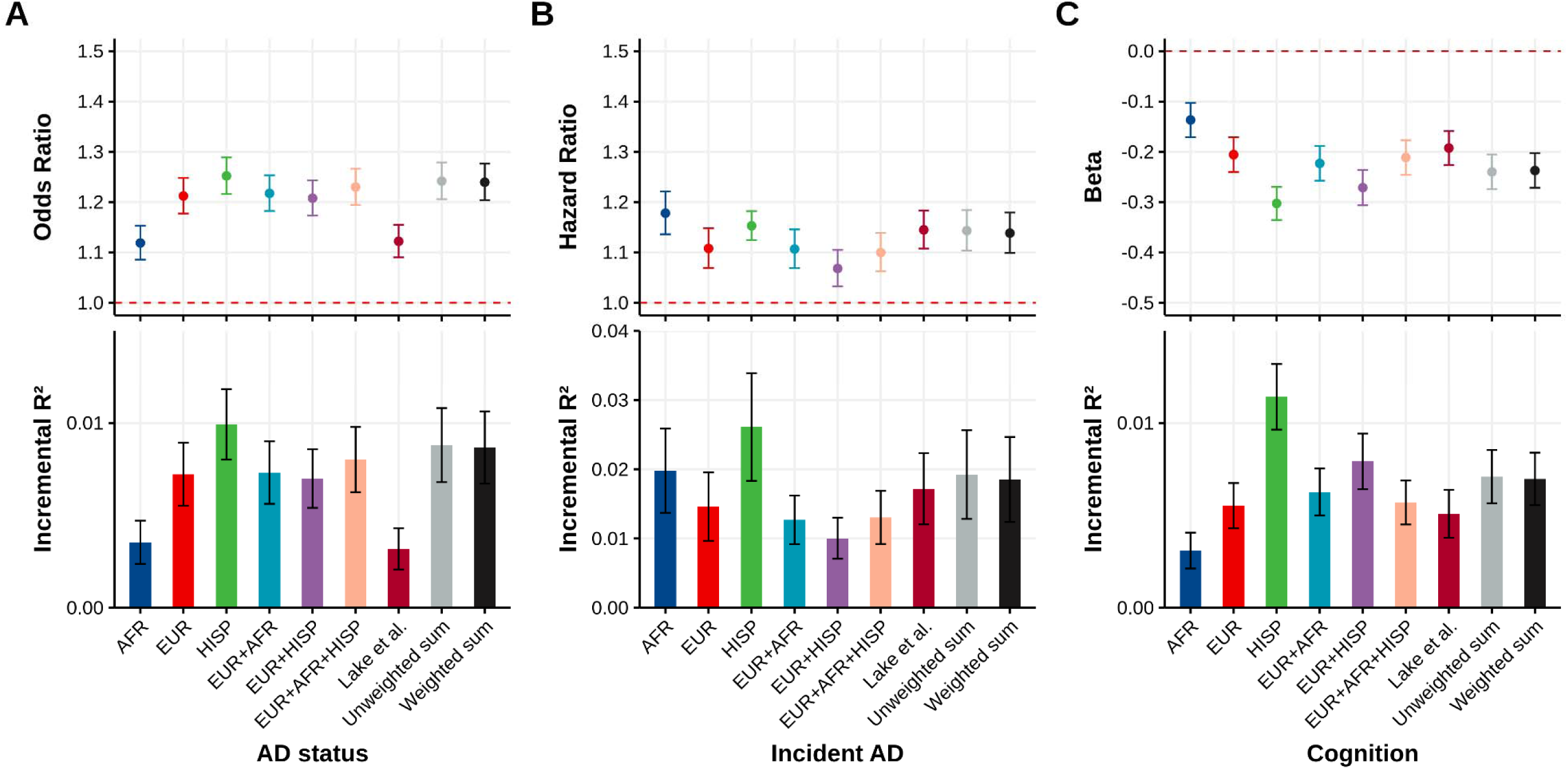
Performance of ancestry-specific, and GWAS-focused multi-ancestry PRS in predicting clinical AD, incident AD, and cognition among Hispanics. Figure 2 presents the mean performance of single-ancestry and multi-ancestry PRS derived from the GWAS-focused multi-ancestry approach, integrating GWAS from African, European, and Hispanic populations, using the clumping and thresholding method, in predicting clinical AD (A), incident AD (B), and cognition (C). Within each panel, each point with an error bar represents the pooled effect size of the corresponding PRS based on 10-repeated 5-fold cross-validation with 95% confidence intervals. Each bar with an error bar represents the mean incremental R² for different PRSs, also based on 10-repeated 5-fold cross-validation with 95% confidence intervals. The performance of each PRS within each fold and repeat for clinical AD, incident AD, and cognition was evaluated using logistic regression, Cox proportional hazards models, and linear models, respectively, with age, sex, cohort, and the first 10 principal components as covariates. ‘EUR+AFR,’ ‘EUR+HISP,’ and ‘EUR+AFR+HISP’ represent in-house meta-analyses combining GWAS from different ancestry groups. ‘Unweighted sum’ and ‘weighted sum’ refer to a simple sum of single-ancestry PRS and a linear combination of each single-ancestry PRS, respectively. All continuous genetic predictors have been standardized to a mean of 0 and a standard deviation of 1 for ease of comparison.

### Validation using plasma biomarkers

*APOE* showed weak to moderate associations with Aβ42 and Aβ42/40, and strong associations with P-tau181 and P-tau181/Aβ42, explaining 1% to 4% of the variance, with the largest variance observed in P-tau181-related biomarkers (Figure 3). Single-ancestry PRS were associated with Aβ42, P-tau181, Aβ42/40, and P-tau181/Aβ42; however, these associations were generally weak, typically explaining less than 1% of the variance across outcomes. Most of these associations were found in PRS constructed from Hispanic and European GWASs, with the strength and presence of these associations varying depending on the PRS method used.

**Figure 3.**
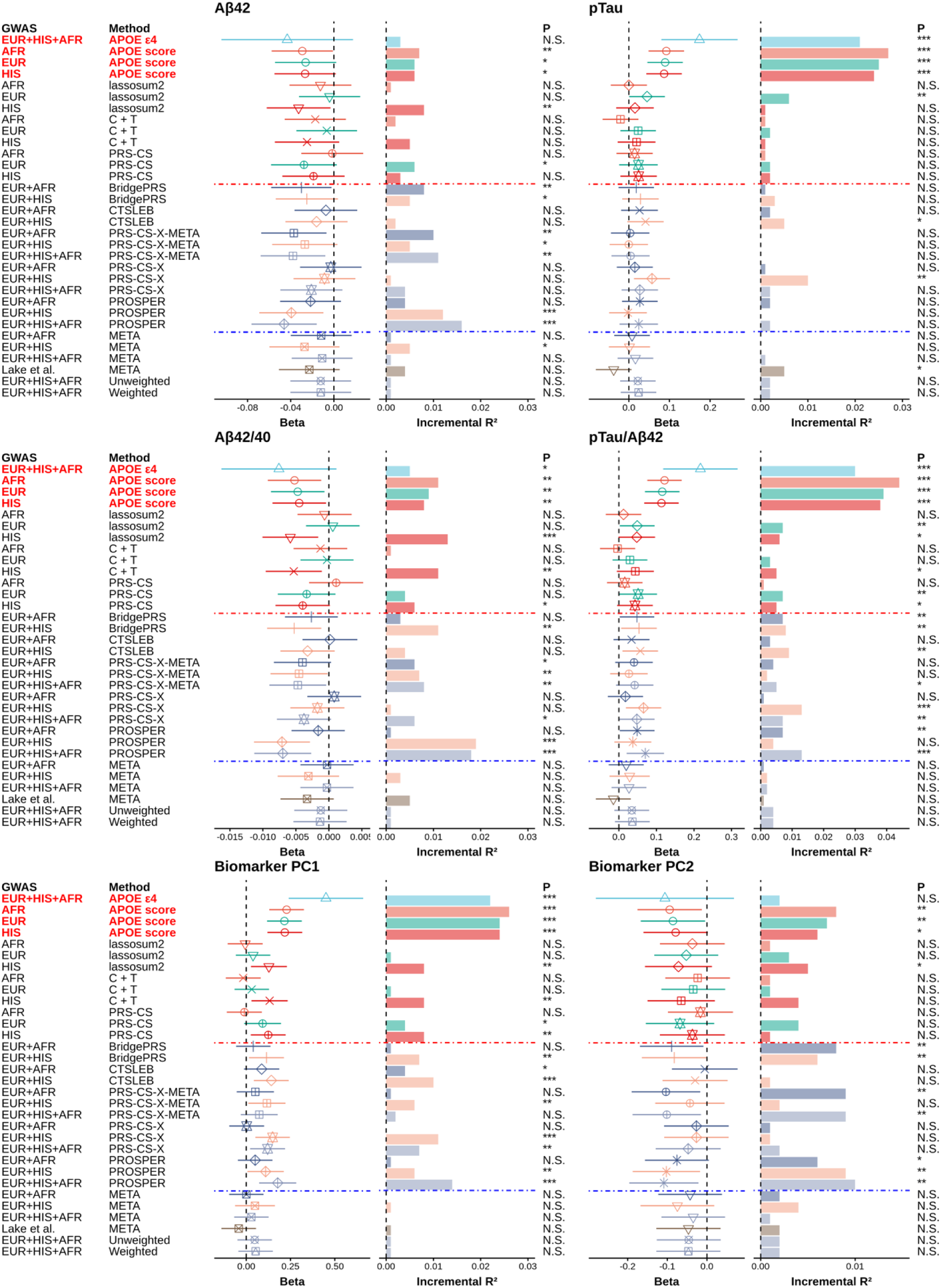
Performance of *APOE*, ancestry-specific, ‘method’-focused and GWAS-focused multi-ancestry PRS in predicting individual and combined biomarkers among Hispanics. Figure 3 represents the prediction performance of *APOE*, single-ancestry, and multi-ancestry PRS derived from both method-focused and GWAS-focused multi-ancestry approaches, integrating GWAS from African, European, and Hispanic populations, in predicting single and combined biomarkers. Within each panel, each point with an error bar represents the beta effect size of the corresponding PRS with 95% confidence intervals. Each bar represents the incremental R² for PRS constructed based on different approaches. The performance of each PRS for each biomarker was evaluated using linear regression, with age, sex, cohort, and the first 10 principal components as covariates. The *APOE* score for each ancestry was weighted by the corresponding *APOE* genotype effect size based on a recent large overview of *APOE* effects across ancestries. The combined effect of biomarkers was constructed using PCA based on four plasma biomarkers—Aβ42/40, P-tau181/Aβ42, NfL, and GFAP—and used the first two PCs as indicators for combined biomarkers. NfL, GFAP, and P-tau181/Aβ42 were the primary contributors to PC1 (loadings ranging from 0.65 to 0.82), all exhibiting positive loadings. For PC2, Aβ42/40 and P-tau181/Aβ42 were the dominant biomarkers with the largest loadings, with Aβ42/40 having the largest positive loading (0.87) and P-tau181/Aβ42 a negative loading (-0.55). Based on the loading direction, a higher PC1 score suggests an increased likelihood of neuronal injury, neuroinflammation, and neurodegenerative profiles, while a higher PC2 score indicates a lower likelihood of AD-specific pathological changes. All continuous genetic predictors have been standardized to a mean of 0 and a standard deviation of 1 for ease of comparison. The results above the red line show single-ancestry PRS, while those below represent ‘Method’-focused multi-ancestry PRS. Similarly, above the blue line are ‘Method’-focused multi-ancestry PRS, and below are ‘GWAS’-focused multi-ancestry PRS. *\*\*\*p < 0.01, **p < 0.05, *p < 0.1, N.S.: non-significant*.

PRS constructed using method-focused multi-ancestry approaches generally outperformed single-ancestry PRS in variance explained across most biomarkers, though the degree of improvement varied across biomarkers. For example, PRS constructed by PROSPER, which integrates European, African, and Hispanic GWASs, consistently showed higher ΔR² values for Aβ42-related biomarkers (e.g., ∼1.6% for Aβ42 and 2% for Aβ42/40). PRS-CSx and PRS-CSx-meta also improved predictions for Aβ42, P-tau181, and P-tau181/Aβ42, though the extent of improvement depended on the biomarker and GWAS involved. Similarly, BridgePRS and CTSLEB outperformed the best single-ancestry PRS for at least one biomarker. In contrast, PRS from GWAS-focused multi-ancestry methods and the simple/weighted sum of the single-ancestry PRS generally performed worse, with no associations detected for most biomarkers.

For combined biomarker analysis, *APOE* was associated with both PC1 and PC2, with the strongest association observed with PC1, explaining about 2.5% of the variance. The *APOE* score explained more variance and showed a stronger association than the *APOE*-ε4 indicator for predicting PC2. For single-ancestry PRS, significant associations were found between PRS based on Hispanic GWAS and PC1 across all methods, but no significant association with PC2. Method-focused multi-ancestry PRS generally outperformed single-ancestry PRS. For PC1, at least one PRS from PRS-CSx, CTSLEB, and PROSPER outperformed the “best” single-ancestry PRS, with PROSPER, integrating all three ancestries, showing the best prediction. For PC2, at least one PRS from BridgePRS, PRS-CSx-meta, and PROSPER outperformed the “best” single-ancestry PRS. PROSPER, integrating all ancestry groups, still showed the best prediction, explaining about 1% of the variance in PC2. Conversely, PRS from GWAS-focused multi-ancestry methods and the simple/weighted sum of single-ancestry PRSs generally performed worse, with no associations detected for either PC1 or PC2.

## Discussion

In this study, we examined the predictive performance of *APOE*, single-ancestry, and multi-ancestry PRSs constructed using method- and GWAS-focused approaches to predict clinical AD, incident AD, cognitive function, and plasma AD-biomarkers among Hispanics. We demonstrated that integrating novel multi-ancestry PRS methods with GWAS across ancestries resulted in improved prediction accuracy than using a single-ancestry PRS constructed based on a European GWAS or a lower-powered African or Hispanic GWAS. The method-focused multi-ancestry PRS also outperformed the PRS constructed from meta-analyses of GWASs across ancestries. To our knowledge, this is the first study that comprehensively evaluated the performance of the PRS based on existing PRS methods and the largest-to-date GWAS for European, African, and Hispanic populations in AD prediction among Hispanics.

The PRS has been extensively studied over the past decade in European populations, proving effective in predicting AD risk^1,23,27^. However, Hispanics remain an understudied due to the lack of adequately sized Hispanic GWAS data and independent cohorts for validation. Two studies reported large PRS effects (OR: 1.38-1.51,) after the *APOE* region was excluded, but these estimates are likely overestimated due to the inclusion of diverse samples and lack of validation^4,5^. In a replication analysis, we adjusted for cohort and family relatedness, which reduced the OR to 1.24 and the ΔR² to 0.8%, aligning with current estimates. Another study involved testing PRS performance in a three-way admixed Colombian population using an internal Hispanic GWAS, finding that the best PRS explained about 0.2% of the variance without *APOE*^7^.

The large variation in PRS prediction performance across studies highlights the importance of identifying the “unbiased” performance of PRS among Hispanics, which we addressed through two internal validation approaches, given the lack of careful validation in past studies. Additionally, studies evaluating the PRS derived from African or large-scale European GWAS in Hispanic populations have shown similarly diminished performance, yielding comparable or worse accuracy than smaller Hispanic GWAS, consistent with our findings^4^. This PRS trans-ancestry portability issue stems from differences in LD, allele frequency, and gene-environment interactions that affect causal-effect sizes^28^.

Despite constructing PRSs for Hispanics using single-ancestry GWAS, strong predictions can also be achieved with diverse GWAS data across multiple ancestries because true causal variants have globally correlated effect sizes shared across ancestry groups^29,30^. Building on this, two strategies have emerged for multi-ancestry PRS: deriving PRSs from multi-ancestry GWAS meta-analyses (GWAS-focused) and using methods that leverage diverse ancestry data (method-focused). In this study, the multi-ancestry PRS from the GWAS-focused approach generally performed worse than the method-focused approach and the PRS derived from Hispanic GWAS in predicting AD outcomes. This aligns with recent findings that multi-ancestry PRSs performed as similar or worse in predicting AD phenotype among Hispanics than single-ancestry PRSs^6,7^. Although multi-ancestry GWAS may produce PRSs that perform uniformly across groups, they do not fully account for LD and effect size differences, making them less optimal for specific groups like Hispanics^9,11,31^. Although combining single-ancestry PRSs can improve predictions for Hispanics, we found that these combined PRSs, although better than those from European or African GWAS, were still less effective than those derived from Hispanic GWAS^6,10^.

The method-focused approach produced the most effective multi-ancestry PRS for predicting AD outcomes among Hispanics, particularly when GWAS data from African, European, and Hispanic populations was integrated. The three-ancestry PRS based on PROSPER or PRS-CSx explained about 1.3-1.6% of the variance in clinical AD, without including *APOE*, which is similar to the PRS performance observed in a recent European study using data from the Alzheimer’s Disease Genetic Consortium^32^. Similar findings were seen for incident AD, cognition, and biomarkers, aligning with expectations given the admixed nature of Hispanics. Across methods, the PRS based on all three ancestries performed as well as or better than those based on the European and Hispanic combination whereas the PRS derived from the European and African combination performed relatively worse because the method-focused approach is intended to maximize prediction in the target sample by combining target population GWAS with information “borrowed” from auxiliary GWAS, making accurate specification of the target population GWAS (in this case, Hispanics, not Africans) essential for achieving the most powerful PRS^8,10,11^. However, in the absence of a Hispanic GWAS (which many researchers lack), some methods (e.g., PRS-CSx/BridgePRS) that integrate GWASs from European and African populations, though not optimal, can still produce a relatively more powerful PRS among Hispanics than any single-ancestry PRS derived from European or African GWASs.

*APOE* consistently shows an association with AD, cognition, and plasma biomarkers, but its effect is notably weaker among Caribbean Hispanics than among non-Hispanic whites. *APOE*-ε4 explained about 1.4% of the variance in clinical AD, with an OR of 1.83, consistent with recent findings^3,33^. For incident AD and cognition, the *APOE* indicator explained about 2% and 1.5% of the variance, respectively. Notably, the best non-*APOE* PRS in this study explained a similar or slightly higher amount of variance across AD outcomes than *APOE*, contrasting with findings among Europeans in which *APOE* effects are substantially greater than the PRS^23^. One possible explanation from previous studies is that this attenuated *APOE* effect may result from the reduced impact of *APOE*-ε4 homozygosity in individuals with greater Amerindian ancestry and the diminished influence of *APOE*-ε4 in those with greater African ancestry^3,34^. The comparable performance of the PRS in this study, relative to *APOE*, highlights the importance of non-*APOE* variants in contributing to AD risk among Hispanics and suggests the PRS could become a more efficient tool for risk profiling in this population, especially with access to more powerful Hispanic or African GWASs in the future.

The PRS derived here shows potential in predicting early changes in plasma biomarkers, consistent with findings in European populations^35,36^. However, an interesting observation is that the PRS and *APOE* seem to capture distinct aspects of AD pathology in this population, as reflected in the single- and combined-biomarker analyses, *APOE* accounts for more variation in P-tau181 levels whereas the PRS explains more variation in amyloid pathology. These findings apply to Hispanic-specific and multi-ancestry PRSs. The observation that *APOE* accounts for more variation in P-Tau levels is consistent with recent findings that *APOE* has better predictive value than the PRS in predicting P-Tau levels^35,36^. However, evidence is mixed for plasma amyloid levels. Although past research on Europeans indicates that *APOE* is a more important predictor in Aβ deposition than PRS, some recent studies report that PRS excluding *APOE* is also significantly associated with amyloid status, with one study reporting that the PRS explains a larger fraction of the variation in amyloid levels than *APOE*-ε4^37,38^.

Our study has several limitations. First, we relied on internal validation rather than independent datasets, which may limit generalizability. However, this approach was sufficient for comparing mean performance across methods rather than identifying a single "best" PRS. Second, the GWAS data for Hispanics and Africans were limited, and better-powered GWASs combined with advanced methods may yield more predictive PRSs. Third, the small biomarker sample size may have impacted the detection of significant associations. Fourth, we focused on selected PRS methods and validated them only among Hispanics, leaving room for the exploration of other methods. Fifth, some participants in the current analyses were also included in Lake et al.^7^, so the PRS derived from Lake et al., might overestimate PRS effects. However, this is not a major concern in the current study, as the PRS derived from Lake et al. performed worse than most other PRSs. Finally, our multi-ancestry PRS was validated only in Caribbean Hispanics, and its performance in other Hispanic or non-Hispanic populations remains unclear. Future research should address these limitations with larger and more diverse cohorts.

The results here demonstrate that integrating novel multi-ancestry PRS methods with GWAS data across ancestries improves prediction accuracy compared to single-ancestry PRSs. This advancement enhances the identification of at-risk individuals for clinical trials and early biomarker screening among Hispanics.

## Data availability statement

The data analyzed in this study is subject to the following licenses/restrictions: the datasets analyzed for this study could be requested through a formal research application to the Washington Heights-Inwood Columbia Aging Project and the Genetic Studies of Alzheimer’s Disease in Caribbean Hispanics (EFIGA). Requests to access these datasets should be directed to https://www.neurology.columbia.edu/research/research-centers-and-programs/alzheimers-disease-research-center-adrc/investigators/investigator-resources.

## Ethics statement

The studies involving humans were approved by the Institutional Review Board of the Columbia University Medical Center. The studies were conducted in accordance with the local legislation and institutional requirements. Subjects from WHICAP and EFIGA have provided signed informed consent before participation.

## Author contributions

YX and RM contributed to the study design. YX, RM, DR, and YG prepared the data and performed the data analysis. YX and RM drafted the manuscript. All authors critically reviewed the manuscript and have approved the final version.

## Acknowledgements

We acknowledge the participants from the Washington Heights-Inwood Columbia Aging Project and the Genetic Studies of Alzheimer’s Disease in Caribbean Hispanics (EFIGA) for their dedication. This study would not be possible without them. We also thank the researchers in WHICAP and EFIGA for their efforts in examining subjects over the years.

