## Supplementary materials for "Identifying Individuals at High Risk for Alzheimer’s Disease Among Hispanics Using Single and Multi-Ancestry Polygenic Risk Scores"

**Supplementary methods**

*Data and phenotype ascertainment/harmonization*

The data analyzed in this study were obtained from individuals of Caribbean Hispanic ancestry enrolled in either the Estudio Familiar de Influencia Genética en Alzheimer (EFIGA) or the Washington Heights-Hamilton Heights-Inwood Columbia Aging Project (WHICAP)^1,2^. EFIGA focuses on individuals of Caribbean Hispanic descent, including those with familial and sporadic late-onset AD, and healthy, non-demented individuals from the Dominican Republic and New York. WHICAP is a community-based, multiethnic study of elderly individuals aged 65 and older, residing in northern Manhattan, New York City.

EFIGA and WHICAP provide detailed information on dementia status, adhering to the same diagnostic criteria and protocols. Specifically, the final diagnosis of dementia in both cohorts was determined using the National Institute on Aging-Alzheimer’s Association (NIA-AA) criteria^3^. This diagnosis was established during diagnostic consensus conferences attended by neurologists, psychiatrists, and neuropsychologists, based on neuropsychological battery results and evidence of impairment in social or occupational functioning. MCI was diagnosed when cognitive impairment was present but did not meet the criteria for dementia, while a cognitively intact diagnosis was assigned when no cognitive impairment was detected or the individual did not meet dementia criteria. Given the same standardized protocols and instruments for phenotype evaluation and assessment in EFIGA and WHICAP, no further phenotype harmonization was performed^4^.

*Plasma biomarkers*

Blood samples were collected via standard venipuncture into dipotassium EDTA tubes^5^. Plasma was prepared by centrifugation at 2,000×g for 15 minutes at 4°C within 2 hours of collection, aliquoted into polypropylene tubes, and stored at −80°C. Plasma biomarker assays were conducted from April to November 2022 using Quanterix Simoa (single molecule array) technology on the HD-X platform (Quanterix, Billerica, MA, USA)^6^. Samples were diluted and assayed in duplicate according to the package insert instructions, utilizing three Quanterix kits: Neurology 3-Plex A (catalog No. 101995) for Aβ42, Aβ40, and T-tau; P-tau181 V2 Advantage (catalog No. 103714) for Tau phosphorylated at threonine 181 (P-tau181); and Neurology 2-Plex B (catalog No. 103520) for NfL and GFAP. Based on existing literature, we chose to focus on Aβ42, Aβ42/Aβ40, P-tau181, and P-tau181/Aβ42 due to their pivotal role in AD.

We evaluated PRS performance and its associations with the combined effect of biomarkers using principal component analyses (PCA) on the correlation matrix of Aβ42/40, P-tau181/Aβ42, NfL, and GFAP. Consistent with previous findings, NfL, GFAP, and P-tau181/Aβ42 were the primary contributors to PC1 (loadings ranging from 0.65 to 0.82), all exhibiting positive loadings^5^. For PC2, Aβ42/40 and P-tau181/Aβ42 were the dominant biomarkers with the largest loadings, with Aβ42/40 having the largest positive loading (0.87) and P-Tau/Aβ42 a negative loading (-0.55). Consequently, a higher PC1 score suggests an increased likelihood of neuronal injury, neuroinflammation, and neurodegenerative profiles whereas a higher PC2 score indicates a lower likelihood of AD-specific pathological changes.

*DNA collection, genotyping, quality control, and genetic principal component*

Genotyped data were cleaned using standard QC procedures in PLINK (v1.9)^7–9^. Briefly, individuals with inconsistencies between self-reported and genetic sex and individuals with genotype missingness > 5% were removed and the SNPs with minor allele frequency (MAF) < 1% and Hardy-Weinberg Equilibrium (HWE) test *p*-value < 10^-06^ and genotype missingness > 5% were removed. All samples were then imputed using the Trans-Omics for Precision Medicine (TOPMed) Imputation Reference panel. After imputation, SNPs with MAF < 1% or a low imputation score (R² < 0.3) were removed. The GRCh38 genome build was used.

We employed the PC-relate method, a model-free approach, to fine-tune ancestry by adjusting for familial relatedness^10^. Initial kinship estimates were obtained using the KING algorithm in the SNPRelate R package with a subset of autosomal genotyped SNPs that were clumped for linkage disequilibrium (LD) using a sliding window of 106 bp and an *r²* threshold of 0.1. PC-AiR was used for PCA to account for variance in population stratification. 1,000 genome samples were utilized to annotate PCs for genetic ancestry. New kinship estimates were computed using the first two PCs to adjust for genetic ancestry, and these updated kinship estimates were used to refine the PCA to account for relatedness. This process was iterated twice to obtain a final set of fine-tuned PCs that account for genetic ancestry adjusted for relatedness along with a final genetic relationship matrix adjusted for genetic ancestry.

*In-house meta-analysis of AD GWAS across ancestries*

We conducted in-house meta-analyses of GWAS data for various population combinations^11–13^, including European with African, European with Hispanic, and a combined analysis of European, African, and Hispanic populations. Association summary statistics from GWAS of different ancestry combinations were aggregated using fixed and random effects models, implemented in PLINK v1.9^7^. A fixed-effects analysis was performed alongside random effects in PLINK, as this approach is commonly used in many GWAS meta-analyses. For the PRS analyses, we focused on the random effects model, as these methods are generally more appropriate for multi-ancestry studies, as previously reported^14^.

*APOE-ε4 status and ancestry-specific APOE score*

*APOE* genotype was first divided into six sub-genotypes (ε2/ε2, ε2/ε3, ε3/ε3, ε2/ε4, ε3/ε4, and ε4/ε4) based on rs7412 and rs429358, and then combined into two groups: *APOE* ε4 carriers (ε2/ε4, ε3/ε4, ε4/ε4) and non-carriers (ε2/ε2, ε2/ε3, ε3/ε3). To generate the ancestry-specific *APOE* score, we applied a natural log (ln) transformation of the OR values reported in the largest-to-date overview of *APOE* genetic risk across ancestries^15,16^. OR values were obtained from Belloy et al., which provided ORs for each *APOE* genotype using ε3/ε3 as the reference. It is important to note that some Hispanic participants in the current analyses are also included in the *APOE* analyses by Belloy et al., so the performance of the Hispanic-specific *APOE* score should be interpreted with caution, as it might overestimate the true *APOE* effects among Hispanics.

*Repeated cross-validation for AD and cognition analysis, and tuning-validation split for biomarker analysis*

For AD and cognition, we conducted ten repeated 5-fold cross-validations to ensure robust performance estimates and minimize overfitting, particularly for methods involving machine learning. We chose ten repetitions of 5-fold cross-validation for computational efficiency because studies have shown minimal improvement beyond 10 repetitions^17^. In each repetition, the data was partitioned differently, with 80% of the sample used for training and 20% for validation. These cross-validations were performed using the ‘caret’ R package, with consistent random seeds to ensure the same individuals were included across all methods.

We employed a tuning-validation split for biomarker analyses because plasma biomarkers were available only for a subset of participants. Parameter tuning was conducted among participants without biomarker data, and each method’s performance was validated using participants with biomarker data. In cognition and biomarker analyses, tuning parameters were initially tested using clinical AD as the outcome and subsequently validated with cognition or biomarkers as the outcome. For all analyses, AUC was reported as the confounder-adjusted AUC for each PRS and was computed using the roc.binary function from the RISCA package^18^. Incremental R² for the Cox model was calculated using the CoxR2 package based on the partial likelihood ratio statistic.

**Supplementary Table 1.** Summary of GWASs Used in PRS Development

| **GWAS name** | **Sample size (cases/controls)** | **Outcome** | **Race/ethnicity** | **Reference** |
| --- | --- | --- | --- | --- |
| Bellenguez et al. | 39,106 cases,  46,828 proxy-AD cases,  401,577 controls | AD and proxy-AD | European | PMID: 35379992 |
| Ray et al. | 2,903 cases,  6,265 controls | AD | African | PMID: 37693582 |
| Qiao et al. | 1,986 cases,  3,124 controls | AD | Hispanic | PMID: 36946865 |
| Lake et al. | 54,233 cases,  46,828 proxy AD cases,  543,127 controls | AD and proxy AD | Multi-ethnic: European, Finnish, East Asian, African, Hispanic | PMID: 37198259 |

**Supplementary Figure 1.** Differences in performance metrics for predicting clinical AD between single-ancestry PRS and multi-ancestry PRS derived from a 'method-focused' approach.

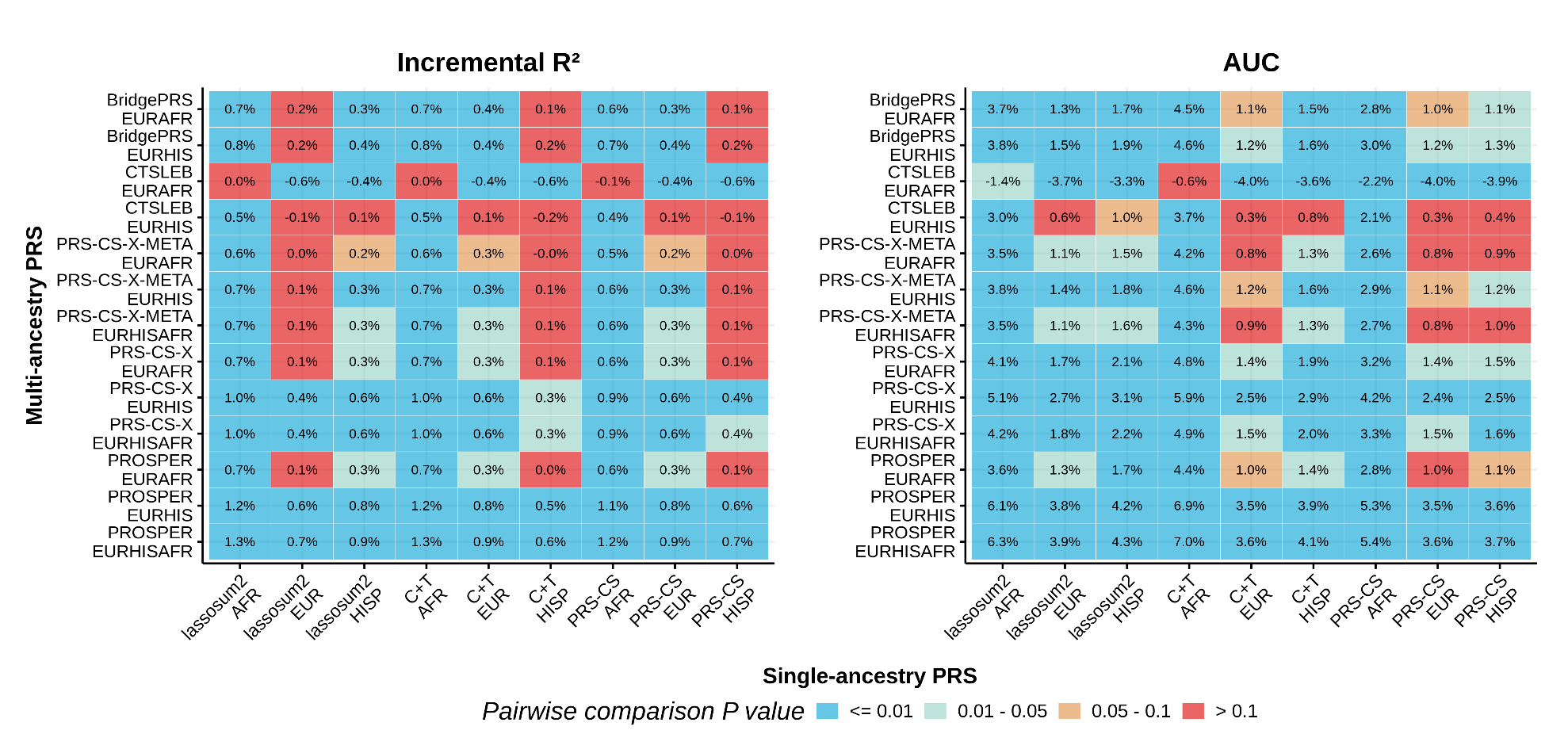

Supplementary Figure 1 presents the mean differences in performance metrics for predicting clinical AD between PRS derived from single ancestry and the 'method-focused' multi-ancestry approach, using a corrected pairwise t-test for cross-validated results. The mean difference is based on ten repeated 5-fold cross-validations. The values within each cell represent the mean difference in prediction accuracy between methods (e.g., Bridge PRS, which integrates European and African GWAS, vs. PRS derived by lassosum2 based on African GWAS). The color represents significance, with annotations provided in the legend.
**Supplementary Figure 2.** Differences in performance metrics for predicting incident AD and cognition between single-ancestry PRS and multi-ancestry PRS derived from a 'method-focused' approach.

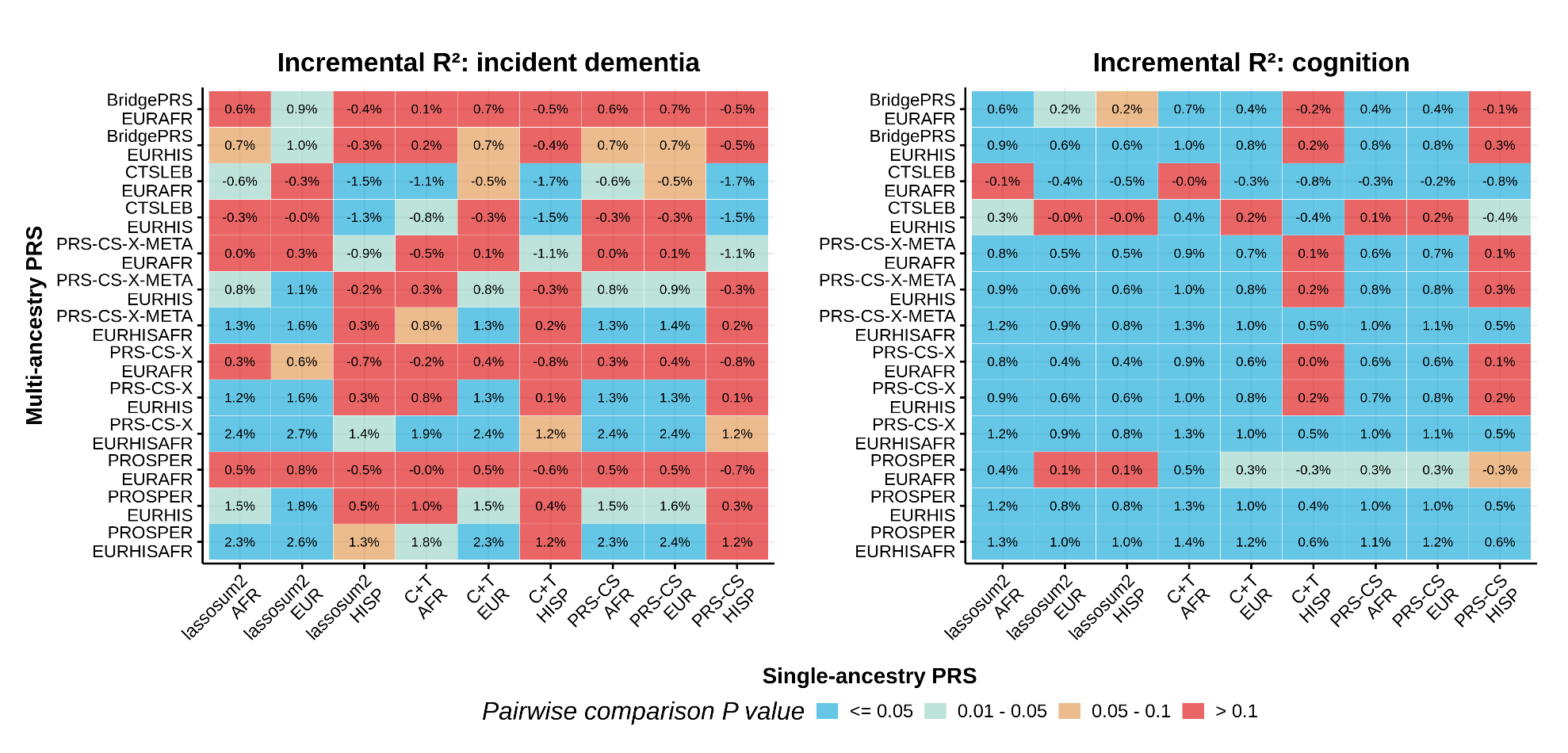

Supplementary Figure 2 presents the mean differences in performance metrics for predicting incident AD and cognition between PRS derived from single ancestry and the 'method-focused' multi-ancestry approach, using a corrected pairwise t-test for cross-validated results. The mean difference is based on ten repeated 5-fold cross-validations. The values within each cell represent the mean difference in prediction accuracy between methods (e.g., Bridge PRS, which integrates European and African GWAS, vs. PRS derived by lassosum2 based on African GWAS). The color represents significance, with annotations provided in the legend.

**Supplementary Figure 3.** Differences in performance metrics for predicting incident AD and cognition between single-ancestry PRS and multi-ancestry PRS derived from a 'GWAS-focused' approach.

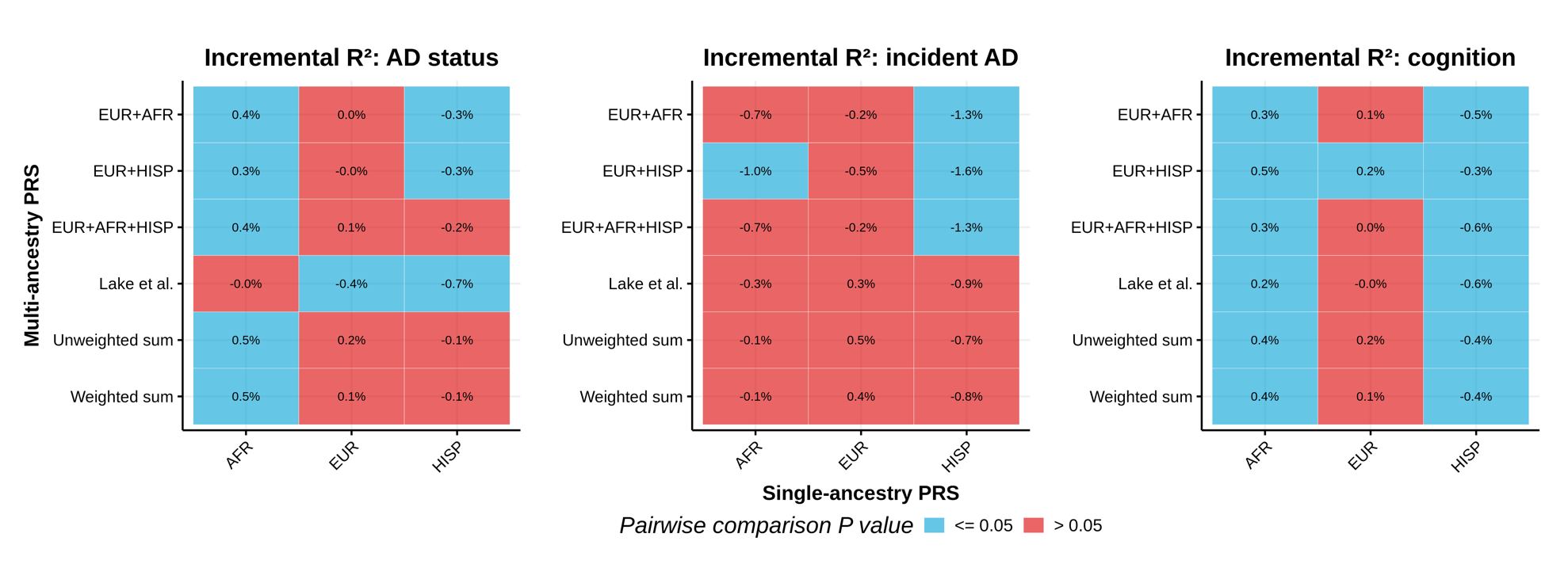

Supplementary Figure 3 presents the mean differences in performance metrics for predicting clinical AD, incident AD, and cognition between PRS derived from single ancestry and the 'GWAS-focused' multi-ancestry approach, using a corrected pairwise t-test for cross-validated results. The mean difference is based on ten repeated 5-fold cross-validations. The values within each cell represent the mean difference in prediction accuracy between methods (e.g., PRS derived from meta-analyses of European and African GWAS, vs. PRS derived from African GWAS). The color represents significance, with annotations provided in the legend. All multi-ancestry PRS derived from the 'GWAS-focused' approach were constructed using the clumping and thresholding method with the target sample as the reference panel.
